# Data Resource Profile: Linking electronic health and social records to study and lower health inequalities in cardiovascular diseases (BIG-HEART)

**DOI:** 10.1101/2025.05.09.25327142

**Authors:** Laura Lõo, Nikita Umov, Marek Oja, Sulev Reisberg, Anneli Uusküla, Raivo Kolde, Taavi Tillmann

## Abstract

The BIG-HEART cohort was established to study and reduce health inequalities in cardiovascular disease by linking rich, multidimensional electronic health and social data across Estonia. The dataset includes all individuals aged 36 and above residing in Estonia in 2012 (N= 770,323). Its full population coverage minimises sampling and healthy volunteer bias. Existing funding and permits will support annual health outcome follow-up through at least 2026, with possible future extensions. The dataset integrates all routinely collected individual-level primary and secondary care health data (including in- and outpatient visits, diagnoses, prescriptions issued and filled), mortality data, and extensive social data (e.g. ethnicity, education, marital status, social benefits, unemployment history, land and business ownership) from eight national registries. This enables exploration of novel social epidemiology dimensions—such as unbiased wealth measures, medication adherence, and care quality—and supports development of equity-enhancing clinical risk prediction algorithms and large language models. Health and social data are linked using pseudonymised identifiers derived from national personal identification numbers, ensuring accuracy and privacy. The data are stored in the OMOP common data model, facilitating international collaboration. Collaboration inquiries are welcome and can be directed to the BIG-HEART team at.

## Data resource basics

### The rationale for establishing BIG-HEART

Cardiovascular diseases (CVD) remain a leading global public health concern despite being largely preventable [1, 2]. 60 years of research has shown a strong association between socioeconomic factors and CVD risk [3]. However, these insights have rarely been integrated into clinical practice. Primary and secondary prevention services can exacerbate health inequalities when attendance at screening programs, preventive medication use, and success with behaviour change services follow social gradients [4–6]. We propose that integrating multidimensional socioeconomic and psychosocial data with electronic health records can first deepen our understanding of the drivers and patterns of health inequity. Second, it can accelerate implementation by enabling developers of risk prediction algorithms and care services to directly incorporate socioeconomic factors into their models and pathways. Existing CVD risk prediction models recommended in clinical practice [7–9] typically incorporate at most one socioeconomic indicator, despite emerging evidence suggesting that each additional layer may enhance both predictive performance and fairness [10–11]. This lack of integration may worsen inequalities from the earliest stage of care—risk prediction— compounded by unequal uptake and adherence to recommended services [12–15]. Estonia has among the highest cardiovascular disease mortality rates in Europe—nearly twice the EU average—and modestly above the global average, which likely reflects higher disease incidence rather than poorer outcomes after diagnosis [16–19]. This underscores both the statistical power and public health potential of conducting CVD research in Estonia. BIG-HEART integrates health and social data to bridge basic epidemiological research with applied solutions, promoting more precise and equitable risk assessments and services.

### Estonian health and social data

Estonia, a country in Northeastern Europe with a population of 1.3 million, has long led globally in building secure and efficient digital infrastructures for public services. Since 1989, every resident has been assigned an 11-digit personal identification code, facilitating data linkage across government systems. In 2001, Estonia launched “X-Road”—a secure data exchange system enabling interoperability across governmental databases while maintaining strict security standards [20]. Managed by the Estonian Information System Authority, this infrastructure supports resilience to changes in underlying database architectures or digital services.

As of 2023, approximately 95% of Estonia’s population is covered by the publicly funded Estonian Health Insurance Fund (EHIF) [21], similar to the UK’s National Health Service [22, 23]. Estonia’s healthcare system includes roughly 400 primary care centers [24] and 20 hospitals [25], each with its own IT system. These local systems are not fully interoperable, allowing flexibility in implementation. However, providers must submit claims, prescriptions, and discharge summaries to the national system, enabling comprehensive, standardised national coverage. Estonia’s national e-prescribing platform records all prescribed medications as well as dispensing status, allowing assessment of medication initiation, adherence, and persistence across population subgroups. Blockchain technology logs all clinician access to personally identifiable health data, enhancing transparency and public trust. Residents can access their records via the national Health Portal [26], similar to systems in Finland and Iceland [27].

### Secondary use of Estonian health data for research

As the collection and coding of electronic health data differ across countries, there is growing interest in harmonising cleaned, research-ready datasets rather than raw data. The Observational Medical Outcomes Partnership (OMOP) Common Data Model (CDM) [28] has become a widely adopted standard for enabling cross-country comparisons and large-scale analyses. Estonia contributed early to this effort, first mapping a 10% random sample of national health data to OMOP [29], followed by a 30% sample that supported several international COVID-19 studies. These analyses provided valuable insights—for example, they confirmed that post-acute COVID-19 symptoms defined by the WHO were frequently reported, though with variation across healthcare settings and countries [30]. They also demonstrated that vaccination, particularly the first dose, significantly reduced the risk of long COVID [31], and showed that serious adverse events like thromboembolism and myocarditis were more common after SARS-CoV-2 infection than in historical cohorts [32].

### BIG-HEART: Advancing health and social data integration

Building on Estonia’s electronic health record infrastructure, BIG-HEART further integrates data by enriching health records with socioeconomic data. BIG-HEART was established in 2024, covering all 770,323 Estonian residents aged 36 and older who as of 01.01.2012 were resident in Estonia according to the Population Register. The cohort is tracked annually for health and mortality outcomes through 2026. Embedded in the OMOP CDM, the cohort supports harmonised, large-scale analyses and international collaborations on health equity.

## Data collected

### Data sources

BIG-HEART is both a longitudinal cohort and a data resource, enabling analysis of health and social exposures over time. It is based on routinely collected, secondary administrative data, structured as a register-based cohort with repeated annual updates. The cohort was constructed using the Population Register, which defined the study population. Table 1 shows baseline demographic characteristics. Data are grouped into (1) health and (2) social domains. Eight national datasets were linked at the individual level via project-specific pseudonyms. Figure 1 provides an overview of the linked data sources, coverage, and age distribution.

**Table 1.**
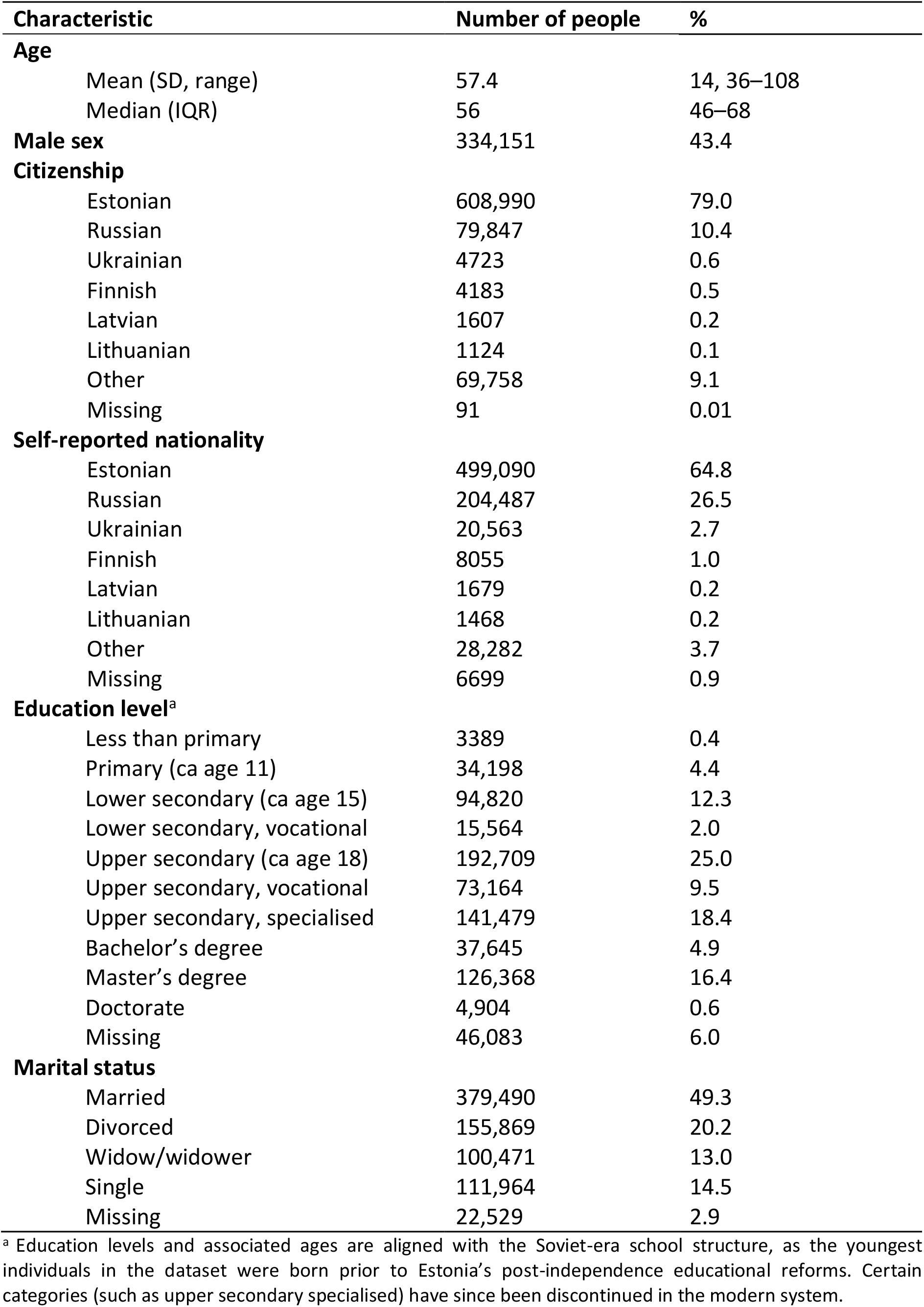
Baseline demographic characteristics of the BIG-HEART cohort (N=770,323) as of 01.01.2012, from the Population Register.

**Figure 1.**
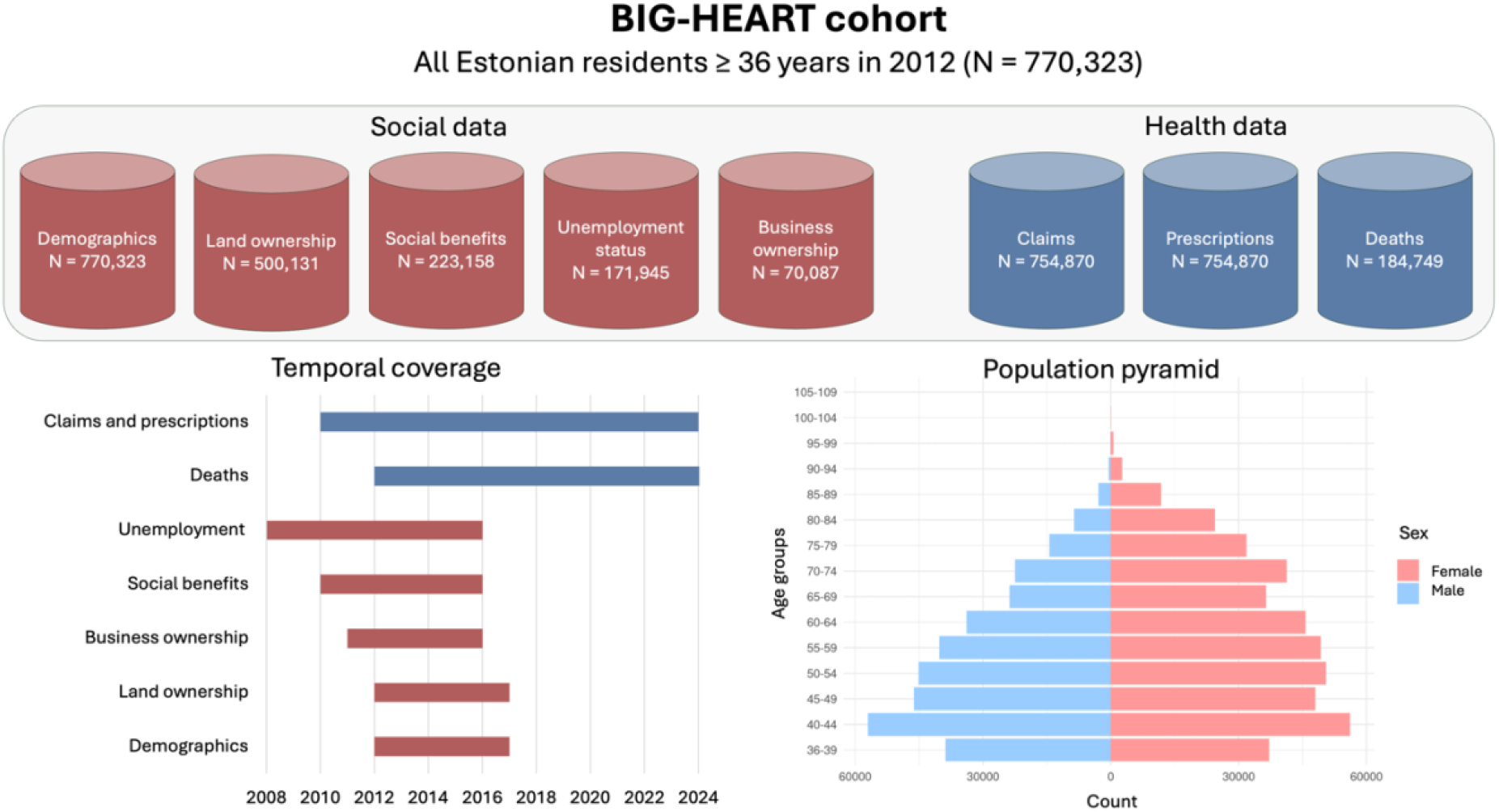
Data sources included in data linkage, data timeliness and population pyramid of the BIG-HEART cohort.

*Health data* are primarily derived from a national billing database managed by the EHIF [33]. This comprehensive dataset includes all reimbursed insurance claims submitted by primary and secondary healthcare providers, covering both in- and outpatient services. Each claim records the procedures performed and diagnoses—both incident and pre-existing—coded according to the International Statistical Classification of Diseases, 10th Revision (ICD-10), and the NOMESCO Classification of Surgical Procedures. Table 2 highlights key diseases of public health relevance, primarily based on the top 30 conditions ranked by Disability Adjusted Life Years (DALY), along with other commonly reported cases from the BIG-HEART database.

**Table 2.** Number of cases of selected diseases with significant public health impact, based on DALY rankings and commonly reported conditions, in the BIG-HEART cohort (N=770,323). Data includes individuals with prevalent disease at baseline or incident disease during follow-up.

| Diagnosis | ICD-10 code | Number of people | % |
| --- | --- | --- | --- |
| Hypertensive heart disease | I10–15 | 487,320 | 63.3 |
| Dorsalgia | M54 | 413,701 | 53.7 |
| Dyslipidaemia | E78 | 327,025 | 42.5 |
| Falls | W00–19 | 314,609 | 40.8 |
| COVID-19 | U07 | 270,460 | 35.1 |
| Virus identified | U07.1 | 176,358 | 22.9 |
| Virus not identified | U07.2 | 139,547 | 18.1 |
| Heart failure | I50 | 188,465 | 24.5 |
| Ischemic heart disease | I21–25 | 136,746 | 17.8 |
| Type 2 diabetes | E11 | 128,924 | 16.7 |
| Cerebrovascular disease | I60–69 | 126,331 | 16.4 |
| Depression | F32–33 | 124,628 | 16.2 |
| Anxiety | F41 | 99,288 | 12.9 |
| Asthma | J45 | 78,040 | 10.1 |
| Chronic obstructive pulmonary disease | J44 | 68,660 | 8.9 |
| Chronic kidney disease | N18 | 63,627 | 8.3 |
| Age-related hearing loss | H91 | 58,593 | 7.6 |
| Prostate cancer | C61 | 22,693 | 2.9 |
| Migraine | G43 | 22,334 | 2.9 |
| Colorectal cancer | C18–21 | 18,119 | 2.4 |
| Breast cancer | C50 | 17,320 | 2.2 |
| Psychotic disorders | F20–25, F28–29 | 14,568 | 1.9 |
| Type 1 diabetes | E10 | 13,621 | 1.8 |
| Lung cancer | C34 | 13,294 | 1.7 |
| Alcohol addiction or use of specialist services | Z71.4, Z72.1, F10 | 12,233 | 1.6 |
| Tobacco addiction or use of cessation services | Z71.6, Z72.0, F17 | 10,519 | 1.4 |
| Other COVID outcomes | U09 | 6204 | 0.8 |
| Pancreatic cancer | C25 | 4692 | 0.6 |
| Use of psychoactive substances | F11–16, F18–19 | 4234 | 0.5 |
| Cervical cancer | C53 | 3696 | 0.5 |
| Alzheimer’s disease | G30 | 3141 | 0.4 |
| Manic episode | F31 | 2189 | 0.3 |
| COVID-19 vaccine causing adverse effects | U12 | 803 | 0.1 |
| Multisystem inflammatory syndrome associated with COVID-19 | U10 | 349 | 0.05 |
| Bipolar affective disorder | F30 | 317 | 0.04 |

The EHIF database contains comprehensive information on sick leave episodes and health insurance coverage, including the start and end dates of coverage and the reasons behind changes in status. Our data suggest that 6–8% of individuals may experience fluctuations in their insurance coverage, implying that stable coverage is closer to 91%, rather than the usually reported 95%. These fluctuations may be due to changes in employment or registration status and should be taken into account when interpreting analyses based on insurance data. In addition to coverage and sick leave data, the EHIF also maintains an extensive prescription records system. This includes detailed information on all dispensed medications—classified using the Anatomical Therapeutic Chemical system—along with the prescribed dosage, prescribing provider, and both the date and location of dispensing. Metadata from packaging codes further enrich these records by identifying the drug manufacturer, strength, and pharmaceutical form (e.g., tablet, cream).

The Causes of Death Register [34] collects comprehensive mortality data. In the BIG-HEART cohort, we have full details on the cause of death when the underlying cause is circulatory (ICD-10 codes I00–I99) or sudden death of unknown cause (R96.0, R96.1). For all other causes, the date of death is recorded, which supports time-to-event analyses and the handling of competing non-cardiovascular outcomes. Should future research require detailed cause-of-death data beyond the circulatory domain, extended access can be requested to support scientifically justified hypotheses.

*Social data* are currently available from five registries. Table 3 presents the socioeconomic and health-related characteristics of the BIG-HEART cohort across the study period. While the primary purpose of collecting social data was to characterise baseline exposures—not to assess the incidence of social events—future extensions may incorporate variables like occupation and income if supported by research hypotheses. Consequently, social data have shorter time coverage than health data. The Population Register [35] includes demographic and migration variables such as sex, birth year, marital status, citizenship, self-reported nationality, county of residence, emigration date, and death date, as well as the most recently recorded educational attainment. Since education data were provided as a 2023 snapshot rather than time-stamped records, we cannot determine participants’ education status at baseline and assume no changes during follow-up.

**Table 3.**
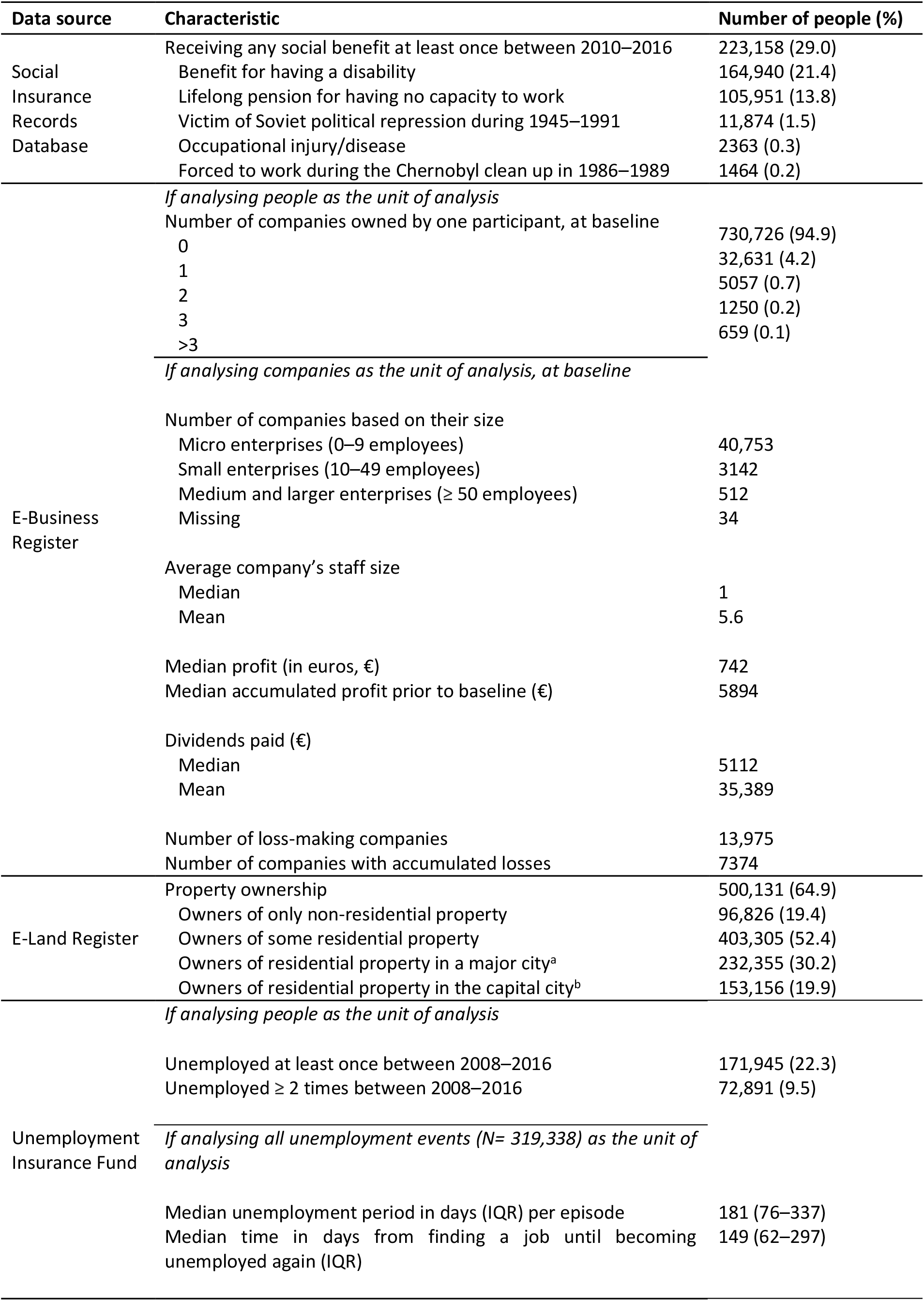
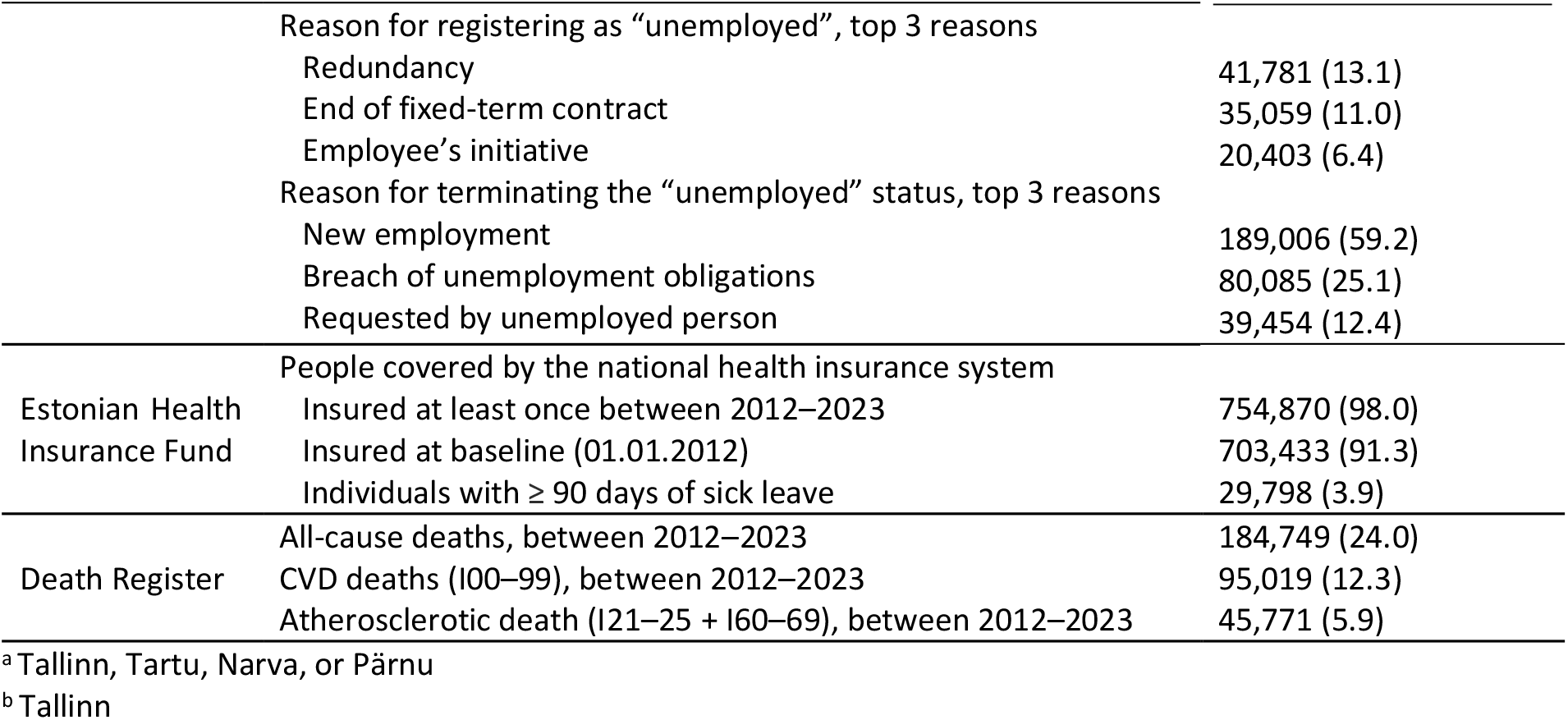
Socioeconomic and health characteristics of the BIG-HEART cohort (N=770,323) over the full study period, from linked databases.

The Social Insurance Records Database [36] covers social benefits – such as type, amount, and payment period – for various reasons, including having a disability, being a victim of severe violence, political repression during the Soviet Union from 1945 to 1991, being forced to work during the Chernobyl clean-up operation from 1986 to 1989, living alone as a pensioner, suffering health damage from an occupational disease or work accident, or receiving an incapacity pension. This data is available from 2010 to 2016.

The Unemployment Insurance Fund data source [37] provides longitudinal data on unemployment episodes, prior occupations, and reasons for entering and exiting unemployment. For the ~25% of participants with at least one unemployment spell, it is possible to reconstruct pre-unemployment income, enabling nuanced analyses of economic vulnerability. Occupations are coded using the International Standard Classification of Occupations, allowing longitudinal tracking of employment stability and career disruption. Evidence suggests that the frequency of labour market transitions may be more predictive of disease incidence than unemployment duration [38]. Among the 10% of participants with multiple unemployment episodes, employment and unemployment often alternate in six-month cycles. The BIG-HEART cohort enables investigation of the contextual and personal factors influencing unemployment resolution—whether through re-employment, retirement, or other paths. Currently, the cohort includes 8 years of employment data, which could be extended to 16 years in future updates if needed.

Objective wealth indicators are available from two registries. The E-Land Register [39] records all landholdings—residential or commercial—owned by participants, down to the neighborhood level (excluding street names). As shown in Table 2, about 65% of the BIG-HEART cohort owns some form of land, enabling rare analyses of how real estate wealth relates to life expectancy, healthcare use, and outcomes—supporting fairness assessments across socioeconomic strata.

The E-Business Register [40] provides data on approximately 40,000 individuals who co-own or operate a legal entity. It offers six years of detailed longitudinal information, including turnover, profitability, tax arrears, employee count, and sector classification using NACE codes. Among the broader population, a smaller share—about 5%—owns business assets, typically through micro-enterprises. This supports research on how financial stability or instability in business ownership affects physical and mental health, healthcare use, and outcomes. Wealth across property and business domains offers a multidimensional view of financial security, precarious employment, and health—especially relevant during external shocks like the COVID-19 pandemic. The dataset could be extended to examine whether business disruptions led to differences in healthcare access, diagnoses, or medication use.

### Secure data processing

To protect privacy, the Population Register assigned a unique, project-specific pseudonym (data key) to each person, replacing national personal identification numbers. These pseudonyms were shared with data providers to enable secure record linkage, but not with the University of Tartu, which never had access to identifiable information. Each data provider used the pseudonym to extract relevant records and returned pseudonymised datasets to a secure computing environment, ensuring that researchers only accessed de-identified data.

The BIG-HEART cohort is stored within one of Estonia’s highest-security computing infrastructures [41], featuring 13,000 CPU cores and 64 GPUs. Approved researchers access the environment remotely via a Virtual Private Network, using a browser-based virtual desktop interface. The system is fully isolated from the public internet. To ensure data protection, copy-pasting is disabled, and all user activity—including video recordings of analytic sessions—is logged for auditing. Data imports and exports are controlled through secure S3 gateways, with every export requiring manual review and approval by a designated principal investigator.

## Data resource use

The BIG-HEART dataset became available to us in the middle of 2024, making it a comparatively new and highly promising resource for studying socioeconomic dimensions of health and healthcare. Although no studies have been published to date, several have been initiated by our comparatively small team to date. Among healthy participants at baseline, we are investigating whether registry-based measures of real estate and business wealth, unemployment episodes, and social benefits predict CVD incidence, and whether such predictors add value to CVD risk prediction algorithms. We are also curious to see if machine learning, artificial intelligence and large language models will create risk prediction models that outperform conventional parametric regression models, both in terms of overall accuracy and algorithm fairness, which the BIG-HEART cohort is exceptionally well placed to evaluate with its rich array of socioeconomic layers and complete population representation. Much of this work is used to inform the design of future interventions, such as identifying persons who are amenable to preventative care (with statins, hypertensives or behaviour change services) and evaluating the effectiveness of novel interventions.

For persons with established disease at baseline, we are describing sociodemographic variations in the uptake and impact of healthcare services, including attendance at screening programmes and medication adherence. We are extending this to also predict which individuals have greater risk of not adhering to medications in the future, and considering pilots to feed this back to frontline clinicians to increase adherence. Beyond these initial opening studies, BIG-HEART is well positioned to address many public health questions that lie at the intersection between computer science, epidemiology, social sciences and the more applied development of novel software as medical devices and related healthcare services. We are keen to collaborate on proposals that either a) analyse the existing data to answer novel questions (which can take as little as 3–6 months to generate results), or b) to add novel data dimensions and layers to the existing dataset via Estonian regulators (which usually takes around 12 months).

## Strengths and weaknesses

BIG-HEART has three core strengths. First, it includes a complete population sample: all Estonian residents aged 36 and above were included via the Population Register. This eliminates sampling bias and sets BIG-HEART apart from biobanks and survey-based cohorts, where participation is voluntary and often limited (5–50% response rates). Second, it offers complete healthcare service capture, due to Estonia’s national electronic infrastructure and OMOP-standardised data. These features support accurate estimation of healthcare use, and of conditions that usually result in clinical contact. Third, BIG-HEART integrates rich social data at the population level. These data, though not originally collected for research, are nearly complete and enable rigorous analysis of social determinants. Social epidemiologists can evaluate how socioeconomic factors affect disease, while data scientists can validate prediction models and assess fairness across subgroups.

Despite these strengths, BIG-HEART has limitations. Some valuable data layers remain unlinked, including the Health Information System (with discharge summaries, clinical notes, and lab results), Estonian Tax Board registries (with declared income and occupation for those never unemployed), and disease-specific registries for myocardial infarction and cancer. Presented health needs may also underestimate true needs. Chronic conditions (e.g. hypertension, obesity, depression) often precede care-seeking and may go undocumented, particularly among disadvantaged groups. This could bias risk estimates and fairness evaluations toward the null. Although official coverage is 95%, baseline data show that only 91% were insured at linkage, and 6–8% cycle in and out of coverage. If uninsured individuals have higher health needs, prevalence and fairness measures may be overly optimistic. Finally, some individuals may have emigrated before baseline or during follow-up without updating their residency. These misclassified participants may be incorrectly considered at risk, biasing estimates toward the null. While we have not yet quantified this, Estonia’s strong incentives for residents to update their status (e.g. for tax, transport, childcare, or mortgage benefits) likely reduce the problem. Future work may identify likely emigrants based on complete disengagement from health and social systems.

## Data resource access

Access to the BIG-HEART data can be obtained with approval from the Research Ethics Committee of the University of Tartu. Researchers can contact the principal investigator Taavi Tillmann through.

## Data Availability

All data produced in the present study are available upon reasonable request to the authors.

## Ethics approval

The BIG-HEART project has received approval from the Research Ethics Committee of the University of Tartu [no. 384/T-8, 20.11.2023].

## Data availability

See Data resource access above.

## Author contributions

T.T. conceived the original research idea and supervised the project. Data analysis was conducted by L.L. and N.U. Data processing and curation were carried out by L.L., N.U., M.O., S.R., and R.K. The original draft of the manuscript was written by L.L., and all authors reviewed, edited, and approved the final version. Funding acquisition was handled by T.T. and R.K.

## Acknowledgements

We acknowledge the contributions of individuals and teams who established the cohorts and data resources that inspired the foundation of BIG-HEART. These include the bespoke CALIBER study of electronic health records (Spiros Denaxas and Harry Hemingway), the

Whitehall II and HAPIEE social epidemiological cohorts (Michael Marmot, Mika Kivimäki, and Martin Bobak), and FinRegistry (Andrea Ganna).

